# The burden of RSV-associated illness in children aged <5 years, South Africa, 2011 to 2016

**DOI:** 10.1101/2022.06.20.22276680

**Authors:** Jocelyn Moyes, Stefano Tempia, Sibongile Walaza, Meredith L. McMorrow, Florette Treurnicht, Nicole Wolter, Anne von Gottberg, Kathleen Kahn, Adam L Cohen, Halima Dawood, Ebrahim Variava, Cheryl Cohen

## Abstract

**Background:** Vaccines and monoclonal antibodies to protect the very young infant against respiratory syncytial virus (RSV)-associated illness are effective for limited time periods. We aimed to estimate age-specific burden to guide implementation strategies and cost effectiveness analyses.

**Methods:** We combined case-based surveillance and ecological data to generate a national estimate of the burden of RSV-associated acute respiratory illness (ARI) and severe acute respiratory illness (SARI) in South African children aged <5 years (2011-2016), including adjustment for attributable fraction. We estimated the RSV burden by month of life in the <1-year age group, by 3-month intervals until 2 years and then 12 monthly intervals to <5 years for medically and non-medically attended illness.

**Results:** We estimated a mean annual total (medically and non-medically attended) of 264,112 (95% Confidence interval (CI) 134,357-437,187) cases of RSV-associated ARI and 96,220 (95% CI 66,470-132,844) cases of RSV-associated SARI (4.7% and 1.7% of the population aged <5 years, respectively). RSV-associated ARI incidence was highest in 2 month-old infants (18,361/100,000 population, 95% CI 9,336-28,466). The highest incidence of RSV-associated SARI was in the <1-month age group 14,674/100,000 (95% CI 11,175-19,645). RSV-associated deaths were highest in the first and second month of life (8.7 deaths/100 000 (95% CI 6.3-11.2) and 8.7 deaths/100 000 (95% CI 6.4-11.4), respectively).

**Conclusion:** Due to the high burden of RSV-associated illness, specifically SARI cases in young infants, maternal vaccination and monoclonal antibody products delivered at birth could prevent significant RSV-associated disease burden.

## Background

Globally, an estimated 3.2 million RSV-associated lower respiratory tract infection (LRTI) hospitalizations and between 60 000 and 118 000 deaths occur annually in children aged <5 years. A large proportion of these hospitalizations and deaths occur in low and middle income countries (LMIC)^1^.

New RSV prevention technologies in the pipeline include maternal vaccination and long acting monoclonal antibodies (MAB)^2^. Describing the burden of RSV-associated mild and severe disease are pivotal for estimating the cost effectiveness of these new interventions. Although incidence data of hospitalization with RSV-associated LRTI from South Africa have been published these data did not include national estimates and did not quantify non-medically attended disease burden nor mild illness associated with RSV^3^. Burden estimates in fine age groups (in infants <1 year) including non-medically illness will also be used to improve the parameterizing of cost-effectiveness models, which will be valuable for policy makers.

Additional burden of disease may lie in mild illness, and therefore leaving this group out of estimates may significantly under estimate the burden, cost and cost effectiveness of interventions^4^. Due to polymerase chain reaction (PCR) testing the identification of viruses in the respiratory tract has become more sensitive, the presence of these viruses may not be associated with disease, therefore adjusting for the attributable fraction of RSV strengthens burden estimates (ref JM AF paper, this paper is submitted). The burden of non-medically attended RSV-associated illness has not been estimated in our setting; however, estimates for influenza suggest that there is significant burden of disease in non-medically attended illness^5^. Although the case fatality ratio for RSV-associated illness is lower than other viral pathogens, such as influenza, estimating the burden is important^6^. More specifically quantifying out-of-hospital deaths in the younger infant is necessary, to complete the burden estimates of RSV-associated illness. Data published in our setting model out of hospital death in the <5, refining these estimates to finer age bands will improve mortality burden^7^. Describing the seasonality of RSV may assist with the implementation of RSV prevention technologies by targeting immunisations and MAB administration prior to and during the peak RSV transmission season^2^.

We aim to describe the full burden of RSV-associated illness in South African children aged <5 years (both medically and non-medically attended illness) in 1-month age groups for infants and 3-month age groups until 2 years and then yearly until <5; specifically, we describe the burden of RSV-associated acute respiratory illness (ARI), severe acute respiratory infection (SARI) and mortality (in- and out-of-hospital) in South Africa during 2011 to 2016.

## Methods

We made use of the modified Fuller method to calculate the burden of medically attended and non-medically attended RSV-associated acute respiratory illness (ARI) and severe acute respiratory illness (SARI) in children aged <5 years in South Africa during 2011 to 2016^8^. Details of the approach and data sources have previously been published^5,9^

### Definitions

For this analysis we defined RSV-associated illness in children aged <5 years by the following case definitions:

1. Medically attended acute respiratory illness (ARI): child presenting to a primary healthcare clinic with influenza-like illness (history or measured fever and cough with duration of symptoms less than 10 days) adjusted to include children with cough <10 days without fever^10^.
2. Non-medically attended ARI: a child with cough with or without fever but who did not present to any healthcare provider/institution excluding pharmacies.
3. Medically attended SARI: a child hospitalized with physician-diagnosed LRTI (including bronchopneumonia, pneumonia, bronchiolitis, bronchitis, pleural effusion) with symptom duration of less than 10 days.
4. Non-medically attended SARI: a child with any one of the symptoms of LRTI (cough, shortness of breath, stridor, chest in-drawing, with or without fever) who did not present to any healthcare provider or institution excluding pharmacies.

We refer to ARI as mild illness and SARI as severe illness in the remaining text.

Since 2009 South Africa has conducted prospective hospital-based sentinel surveillance for severe respiratory illness in five of the nine South African provinces. In 2012, outpatient surveillance was added at clinics in the catchment area of two sentinel hospitals. Sentinel site data from the Edendale Hospital (and Gateway Clinic) in KwaZulu-Natal Province (KZN), and the Klerksdorp-Tshepong Hospital Complex (KTHC) (and Jouberton Clinic), in the North West Province (NWP) supply data for the base provincial estimates^8^. Details of the surveillance methodology are available in the supplementary material.

In-hospital mortality was estimated by applying the age-specific observed (pneumonia surveillance programme) case-fatality ratio to the national number of RSV-associated severe illnesses in this burden analysis. To estimate out-of-hospital mortality we used published data on the proportion of deaths that occurred out-of-hospital. In-hospital deaths were adjusted for this proportion to give the total estimated number of deaths ^11^.

Non-medically attended illness was estimated using data from health utilization surveys conducted in three provinces^12,13^. Risk factor (relative risks) estimates for pneumonia in the additional seven additional provinces, including HIV infection, exposure to indoor air pollution, crowding, malnutrition, low birth weight and non-exclusive breastfeeding were obtained from published data and the DHS^14,15^. Attributable fraction of RSV estimates were obtained from an analysis done comparing controls to individuals with mild illness and controls to individuals with severe illness in our setting (RSV AF paper).

Population denominators were obtained from the National Census of 2011, annual adjustments were made to these estimates using regional and national adjustment factors supplied by Statistics South Africa^16–18^. Population data is supplied in one-year age bands. In the <1-year group, an adjustment factor taking into consideration neonatal and infant mortality rates, described in the demographic health survey, was applied to allocate population by month of life^3^.

### Data analysis

#### Estimating the number and incidence rate of RSV-associated medically attended SARI

##### Step 1: Estimating severe illness rates in base provinces (NWP and KZN)

We estimated RSV-associated severe illness hospitalization rates for Edendale and Klerksdorp hospitals, these were adjusted for non-enrolment due to weekends, refusals/non-enrolment on surveillance days, and health seeking behavior. The population denominators for the hospital catchment area were derived from census projections and rates of severe illness in the catchment area were applied to provincial population denominators. The method for estimation of the adjusted rate of severe illness is described in the supplementary document (ref supplementary document).

##### Step 2: Estimate the severe illness rate in the other 7 provinces

To estimate the SARI hospitalizations in the other 7 provinces the base provinces (NWP and KZN) SARI rates were adjusted for the each of the seven provincial-level prevalence of risk factors for pneumonia including HIV infection, exposure to indoor air pollution, crowding, malnutrition, low birth weight and non-exclusive breastfeeding. (ref supplement)

##### Step 3: Estimation of rates of RSV-associated severe illness in all provinces

The provincial rates of RSV-associated hospitalization were calculated by multiplying the provincial severe illness hospitalization rates by RSV detection rates (number of RSV positive severe illness cases/ number of severe illness tested) obtained from the measured RSV-detection rates at sentinel surveillance sites (all sentinel sites) and adjusted by the attributable fraction of RSV (supplementary document).

##### Step 4: Estimation of the number and rate per 100 000 population of RSV-associated severe illnesses in all provinces

We estimated the provincial number of RSV-associated SARI hospitalizations by multiplying the provincial RSV-associated severe illness rates by the population at risk in each province over the study period. Provincial estimates are added together to obtain a national estimate. National rates are estimated by dividing total illness episodes by the national population and presented per 100,000 persons (supplementary document).

#### Non-medically attended RSV-associated SARI

The same methodology was applied as for hospitalized RSV-associated severe illness making use of non-medically attended numbers and rates. Using the following four steps; (i) estimating the non-medically attended severe illness rates in the base province from a Healthcare Utilization Survey (HUS) conducted in the communities of the base hospital, to describe the proportions of individuals who seek formal or non-formal healthcare for the symptoms of mild or severe illness. ii) Estimate the non-medically attended severe illness rates in other provinces applying the same adjustments for the risk factors for pneumonia as for medically attended severe illness obtained from the DHS. (iii) Estimate the non-medically attended RSV-associated severe illness rate by applying the RSV-detection rate from our surveillance data and (iv) Estimate the national non-medically attended number of RSV-associated severe illnesses by multiplying provincial rates by provincial population estimates and then sum the provincial estimates. National rates are estimated by dividing total illness episodes by the national population and presented per 100,000 persons (supplementary document).

#### Estimating the number and incidence rate of RSV-associated mild illness, non-hospitalized cases

Surveillance for mild illness is only conducted at one of several clinics serving each of the base hospital catchment areas using the case definition of ILI. The base populations are derived by a backwards adjustment of the severe illness rate at sentinel hospitals, adjusting for mild illness cases transferred to hospital and in-referrals for other facilities (ref supplementary). We use mild illness rates at sentinel sites as proxies for the base provinces and expand to other provinces and nationally in a similar way to the severe illness and RSV-associated severe illness estimates but without adjusting for the severe illness risk factors. As RSV-associated illness may frequently present without fever we adjusted our ILI estimates by multiplying ILI estimates by an adjustment factor for the proportion of ARI without fever^10^. This provides the ARI estimate (mild illness).

Confidence intervals (CI) for medically and non-medically attended RSV-associated illness are obtained by using bootstrapping resamples over 100 replications for all parameters. The lower limit and upper limit of the CI are the 2.5^th^ and 97.5^th^ percentiles of the estimated values from 1000 resampled datasets.

#### Seasonality

The seasonality of RSV-associated illness was derived from the ILI and SARI surveillance programmes mentioned above. The weekly number of positive RSV-cases (ILI and SARI) are divided by the total number of cases to define the detection rate of RSV. The detection rate is plotted by epidemiologic week. A mean weekly detection rate over 5 years is used to define the average seasonality.

### Ethics approval

The SARI and SRI protocol was approved by the University of the Witwatersrand Human Research Ethics Committee (HREC) and the University of KwaZulu-Natal Human Biomedical Research Ethics Committee (BREC) protocol numbers M081042 and BF157/08 respectively. The ILI protocol was approved by HREC and BREC protocol numbers M120133 and BF080/12. respectively. The healthcare utilization survey was approved by HREC M120367 and BREC BE209/13. This surveillance was deemed non-research by the US Centers for Disease Control and Prevention (non-research determination number: 2012-6197).

## Results

### National burden of RSV-associated illness (medically and non-medically attended)

The mean annual number of RSV-associated mild illness cases in children aged <5 years was 264,112 (95% confidence interval (CI) 134,357-437,187); of these 36% (96,783) were in children aged <1 year (Supplementary Table 1). The mean annual number of RSV-associated severe illness cases in children aged <5 years was 96,220 (95% CI 66,470-132,844). Eighty-two percent of these, 78,571 (95% CI 56,187-105,831) were in children aged <1 year (Supplementary Table 2).

### National burden of RSV-associated mild illness

The highest burden of RSV-associated mild illness was in the 2-month age group with a total of 17,259 (95% CI 8,776-26,759) cases or 18,361/100,000 population (95% CI 9,336-28,466). This decreases to 4,873/100,000 (95% CI 1,976-8,259) by 11 months but increases again to 11,935/100,000 (95% CI 6,952-1,8025) in the 12-14-month age group and to 10,261 (95% CI 4,305-17,507) in the 21-23-month age group. (Supplementary table 1). The burden of illness is higher for non-medically attended cases in all age groups (Figure 1).

**Figure 1:**
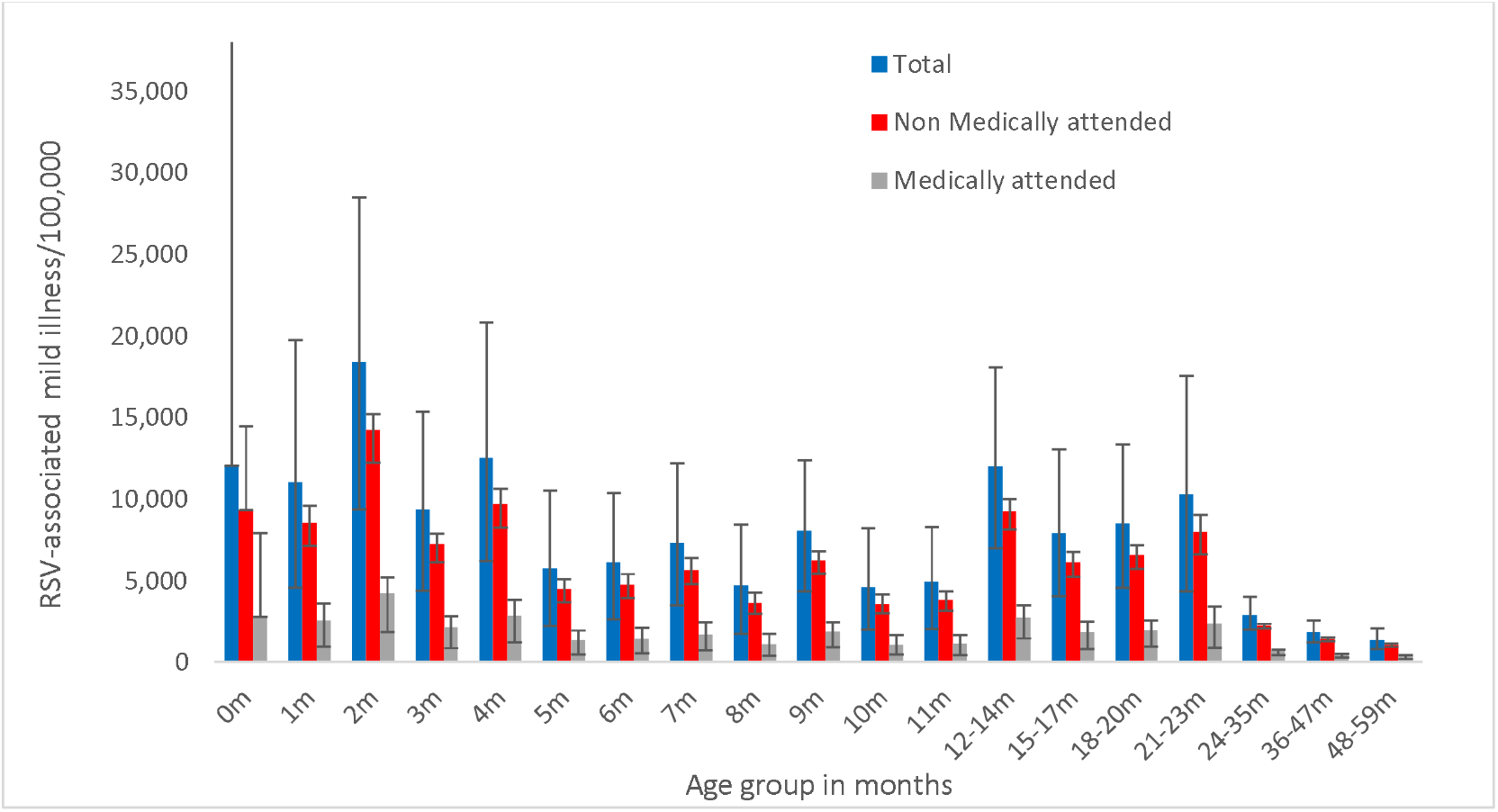
Burden of RSV-associated mild illness, medically and non-medically attended by age group, South Africa 2011-2016. Error bars indicate 95% confidence intervals.

### National burden of RSV-associated severe illness

The highest burden of RSV-associated severe illness was in the <1-month age group, with 14,110 (95% CI 9,784-18,889) cases nationally. Thirty-six percent of cases in children aged<1 year are in the <1 month and 1-month age groups (27,982/78,571). The rate per 100,000 was similar in the <1 month and 1-month age groups (14,674 95% CI 10,175-19,645 vs 14,736 95% CI 11689-18472). The rate of RSV-associated SARI decreased to 2,156/100,000 (95% CI 1,192-3,439) by 11 months. Medically attended cases accounted for 54% (51,605/96,220) of severe illness cases in children aged <5 years (Supplementary table 2). Across all age groups the number of medically attended severe cases is higher than non-medically attended severe cases (Figure 2).

**Figure 2:**
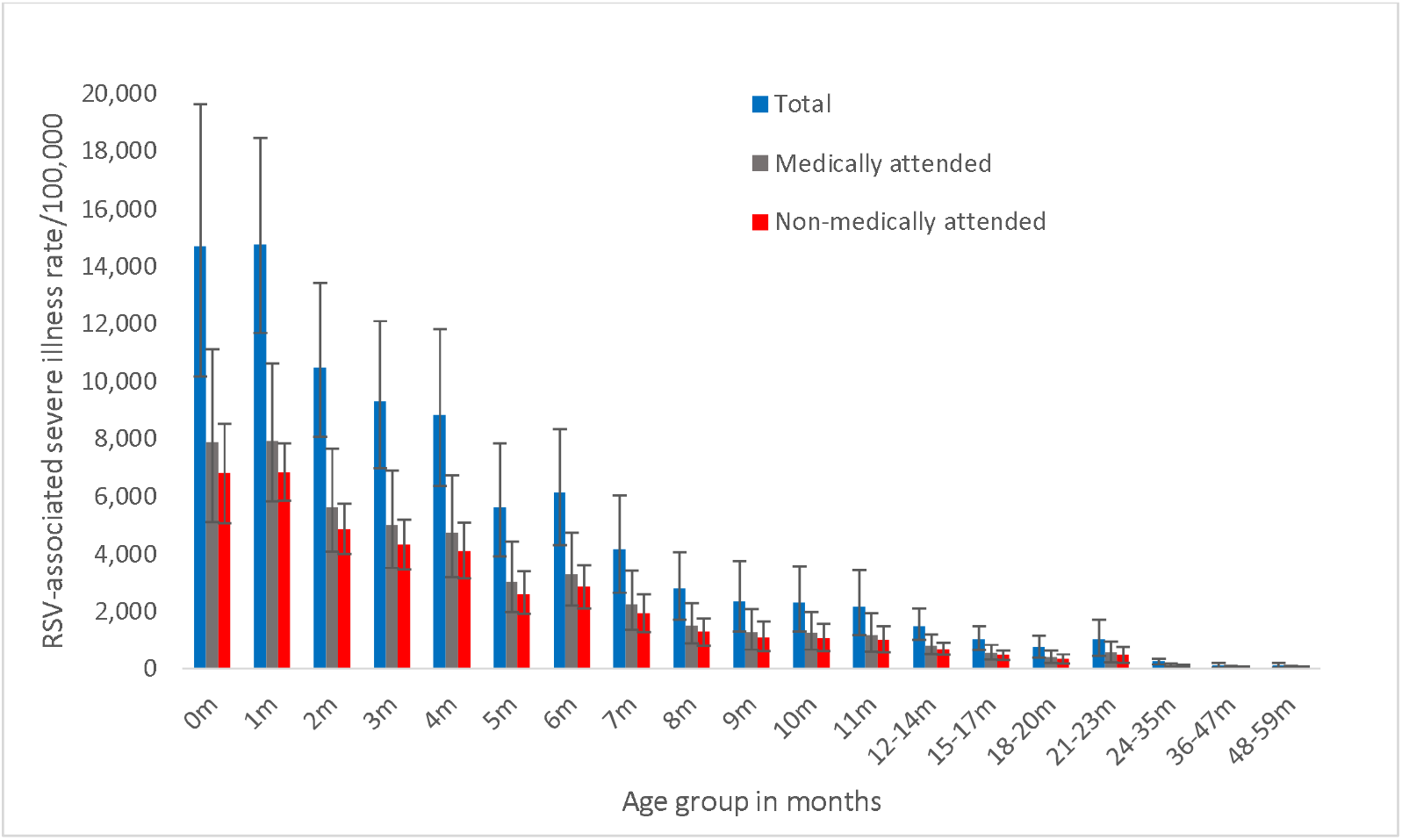
Burden of RSV-associated severe illness, medically and non-medically attended by age group, South Africa 2011-2016. Error bars indicate 95% confidence intervals.

### RSV-associated mortality

The rate of RSV-associated deaths was highest in the first and second month of life 8.7 deaths/100,000 (95% CI 6.3-11.2) and 8.7 (95% CI 6.4-11.4), respectively. Out of hospital death accounted for 26% of the total deaths in children aged <5 years.

### Seasonality of RSV

Although RSV is detected throughout the year in South Africa, an annual peak of RSV circulation is observed in the months of April and May. The mean peak of RSV circulation (2011-2016) was week 14 (second week of April), with a mean peak detection rate of RSV in children hospitalized with LRTI of 42% (range 39% to 65%) (Figure 3).

**Figure 3:**
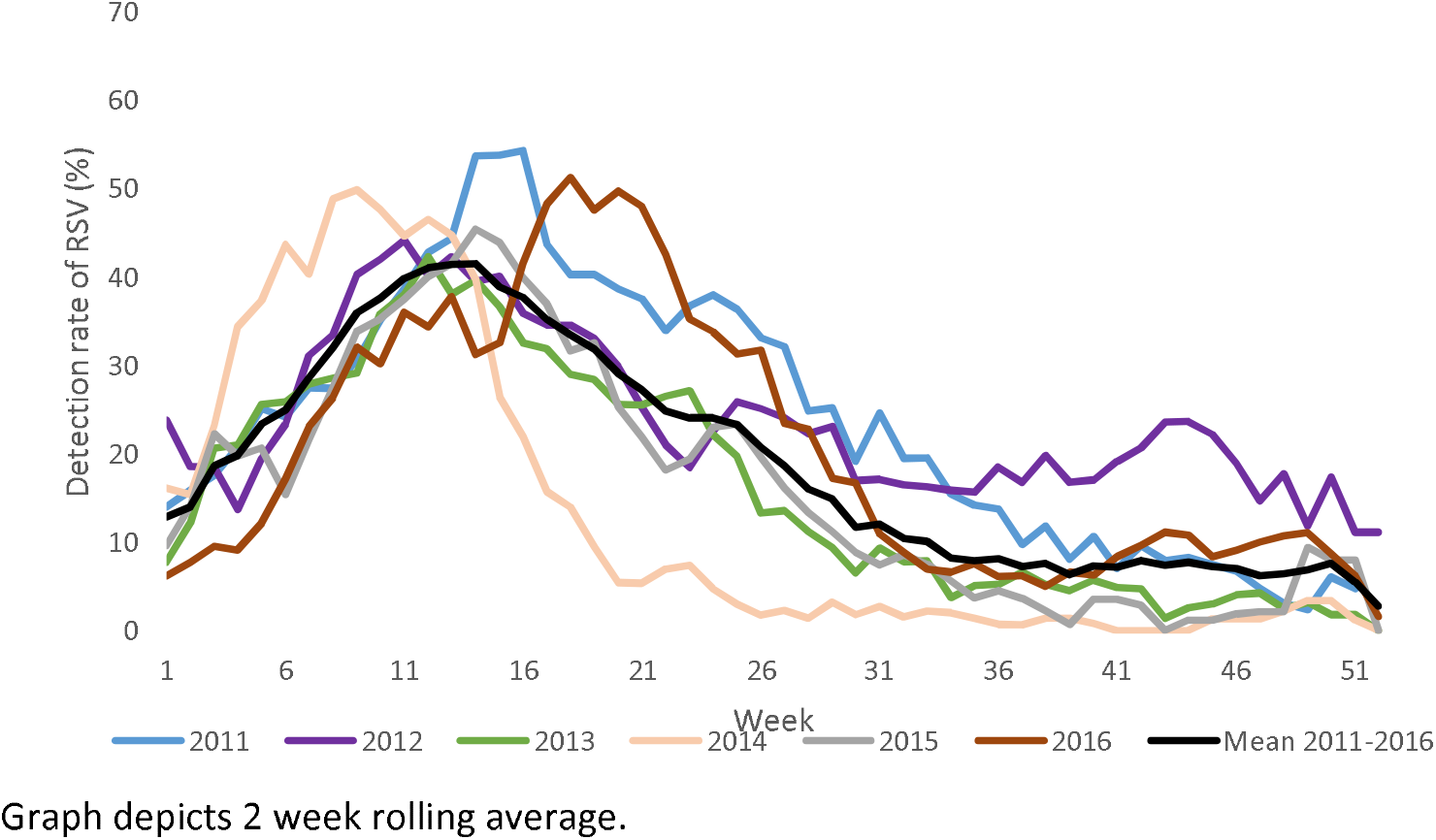
Weekly detection rate of RSV-associated severe illness 2011-2016, in children aged <5 years, South Africa 2011-2016.

## Discussion

We were able to document the national burden of RSV-associated illness in young infants by month of life for both mild and severe RSV-associated illness in a LMIC, specifically demonstrating the highest burden of illness is in the first and second month of life. This analysis also describes the significant burden of non-medially attended RSV-associated illness and is the first to document this burden on our setting. This more inclusive description of burden supports interventions such as maternal vaccination and long acting MABs which target the younger infant. Our analysis adds methodologic techniques that provide more useful data on the burden of RSV in South Africa: Estimating medically and non-medically attended illness in each of these narrow age bands, adjusting estimates for RSV cases without fever and using RSV-AFs to adjust the observed proportion of cases for a more robust estimate.

The burden of disease for mild illness is highest in the 2-month age group and the burden of non-medically attended mild illness is higher than medically attended mild illness in all age groups. We adjusted RSV-associated ILI for the proportion of children who present with RSV-associated illness without fever. Nyiro at al (2018) describe that up to 25% of children with RSV-associated mild illness present without fever^10^. While several estimates of RSV-associated mild or outpatient illness have been published from LMIC, few provide data in fine age bands. In Kenya, Emukule et al (2014) described the outpatient burden of RSV-associated ARI including an estimate of non-medically attended RSV. The incidence of non-medically attend RSV-associated ILI (6,0/1,000 (95% CI 5,4-6,5) in the <5-year age group was higher compared to 4,7/1,000 (95% CI 3,3-66) for medically attended ILI^4^, these estimates are similar to our estimates in this broad age group (<5 years) but do not allow for comparison in finer age bands.

The burden of RSV-associated severe illness is highest in the first month and second month of life after which the burden declines to 11 months of life and further through years 2 to 4 of life. Similar to the severe RSV-associated illness burden, mortality due to RSV-associated respiratory illness is highest in the first 3 months of life, declining steadily until 11 months of life, with few deaths in the older age groups. There are data on the burden of RSV-associated severe illness in other LMIC, but most of these are in wider age bands than what we present here. Shi et al (year) published a meta-analysis of global RSV-associated hospitalization incidence; these results reflect large burden of disease in infants less than 4 months of age in LMIC countries (186/1,000, 95% CI 98-355) and are similar to our estimates in the same age group ^1^. However, our data estimates the burden in <1-month, <2-month and <3-month age groups, illustrating how larger age bands dilute the substantial burden in the first two months of life. In Kenya the burden of disease for severe cases included non-medically attended, for severe RSV-associated illness in children aged <1 year were 14,5/1000 (95% CI 8,9-23,7) which is lower than our estimates of 3,752/100,000 (95% CI 2,530-5,357). However, our estimates for non-medically attended mild illness (6,657/100,00 (95% CI 2692-12540)) were higher than the estimates for mild non-medically attended illness in Kenya 8,9/1000 (95% CI 4,8-16,7). This may be due to a number of factors including socioeconomic factors and access to care.

An additional strength of our study was the ability to adjust numbers by the AF-RSV. This is important because the detection of respiratory viruses does not always reflect their role in causing disease ^19^ (and my af paper). While the AF for RSV was generally high in infants, in children aged 1-4 years the AF-RSV was below 85% for example 74.6% (95% CI 53,6%-86,0%) for mild illness and 83,4% (95% CI 70,9%-90,5%) for severe illness.

Out-of-hospital mortality is difficult to define, specifically where cause of death is not documented. In an earlier analysis of excess mortality attributable to RSV, we estimated that approximately 26% of RSV-associated deaths in children aged <5 years occur in the community^11^.That model estimated 665 (95% CI 105-1105) deaths in children aged <5 years to be RSV-associated very similar to this estimate of 650 (95% CI 479-947) deaths in children <5 years. Shi et al estimated that up to 49% of deaths from RSV-associated SARI, in children <5 year, in LMIC countries occur out of hospital^1^. Other estimates from recent publications suggest that out of hospital deaths may account for a larger proportion of RSV deaths for example in India >80% of RSV-associated deaths occur in the community, in Zambia the estimate was 62% and in Pakistan 27%^20,21,22^. These newly published data suggest that our estimate may be a minimum estimate.

Variation in RSV seasonality by country, region and climatic zone is well described, implying that the description of seasonality in each country is important ^23,24,25^. Describing country-specific seasonality will assist with decision making regarding the seasonal vs all year administration of interventions such as maternal vaccine and MAB treatment for young infants. Knowledge of RSV-seasonality will also provide important data for cost-effectiveness models.

Limitations of our study include: Although we were able to make use of observed data in many provinces (5/9), not all provinces of the country were included in the observed data. Population data were adjusted up from the 2011 census and may not accurately account for population movement and increases.

## Conclusion

The high burden of RSV-associated illness in children in South Africa, particularly very young infants, places significant burden on healthcare systems. Interventions such as maternal vaccines and long acting monoclonal antibodies may have a substantial impact on the burden to the healthcare system and the health of infants.

## Data Availability

All data produced in the present study are available upon reasonable request to the authors

---

**Table 1:** Estimated mean annual number and rates of respiratory syncytial virus-associated deaths in children aged <5 years, South Africa. 2011-2016.

| Age group:<br>months |  |  |  |  |  |  |
| --- | --- | --- | --- | --- | --- | --- |
|  | Total |  | Medically-attended deaths<sup>a</sup> |  | Non-medically attended deaths <sup>b</sup> |  |
|  | Number (95% CI) | Rate<sup>c</sup> (95% CI) | Number (95% CI) | Rate<sup>c</sup> (95% CI) | Number (95% CI) | Rate<sup>c</sup> (95% CI) |
| <1 | 109 (73-142) | 110.8 (74.8-144.5) | 81 (56-108) | 82.0 (56.9-109.8) | 28 (18-34) | 28.8 (18.0-34.7) |
| 1 | 107 (83-131) | 111.3 (86.0-135.8) | 79 (63-99) | 82.4 (65.3-103.2) | 28 (20-31) | 28.9 (20.6-32.6) |
| 2 | 76 (57-95) | 79.1 (59.5-98.7) | 56 (43-72) | 58.6 (45.2-75.0) | 20 (14-23) | 20.6 (14.3-23.7) |
| 3 | 67 (49-85) | 70.2 (51.3-89.0) | 50 (37-65) | 51.9 (39.0-67.6) | 17 (12-20) | 18.2 (12.3-21.4) |
| 4 | 64 (45-83) | 66.6 (46.8-86.9) | 47 (34-63) | 49.3 (35.6-66.1) | 17 (11-20) | 17.3 (11.2-20.9) |
| 5 | 40 (27-55) | 42.4 (28.8-57.6) | 30 (21-42) | 31.3 (21.9-43.8) | 11 (7-12) | 11.0 (6.9-13.8) |
| 6 | 40 (27-53) | 41.6 (28.6-55.2) | 29 (21-40) | 30.8 (21.7-41.9) | 10 (7-13) | 10.8 (6.9-13.2) |
| 7 | 27 (17-38) | 28.1 (17.6-39.9) | 20 (13-29) | 20.8 (13.3-30.3) | 7 (4-9) | 7.3 (4.2-9.6) |
| 8 | 18 (11-26) | 18.9 (11.3-26.8) | 13 (8-19) | 14.0 (8.6-20.4) | 5 (3-6) | 4.9 (2.7-6.4) |
| 9 | 15 (8-24) | 16.0 (8.6-24.8) | 11 (6-18) | 11.8 (6.5-18.9) | 4 (2-6) | 4.2 (2.1-6.0) |
| 10 | 15 (8-22) | 15.7 (8.6-23.6) | 11 (6-17) | 11.6 (6.6-17.9) | 4 (2-5) | 4.1 (2.1-5.7) |
| 11 | 14 (7-21) | 14.6 (7.9-22.7) | 10 (6-16) | 10.8 (6.0-17.2) | 4 (2-5) | 3.8 (1.9-5.4) |
| 12-14 | 14 (9-19) | 4.8(3.2-6.7) | 10(7-14) | 3.6 (2.5-5.1) | 4 (2-5) | 1.3 (0.8-1.6) |
| 15-17 | 10 (6-13) | 3.4 (2.1-3.7) | 7 (5-10) | 2.5 (1.6-3.6) | 2 (1-3) | 0.9 (0.5-1.1) |
| 18-20 | 7 (4-10) | 2.5 (1.3-5.5) | 5(3-8) | 1.8 (1.0-2.8) | 2 (1-3) | 0.6 (0.3-0.9) |
| 21-23 | 10 (4-15) | 3.4 (1.5-5.5) | 7 (3-12) | 2.5 (1.2-4.2) | 2 (1-4) | 0.9 (0.4-1.3) |
| 24-35 | 9 (6-13) | 0.8 (0.5-1.2) | 7 (5-10) | 0.6 (0.4-0.9) | 2 (1-3) | 0.2 (0.1-0.3) |
| 36-47 | 4 (2-7) | 0.4 (0.2-0.7) | 3 (2-6) | 0.3 (0.1-0.5) | 1 (0-2) | 0.1 (0.0-0.2) |
| 48-59 | 5 (2-7) | 0.4 (0.2-0.7) | 3 (2-6) | 0.3 (0.1-0.5) | 1(0-2) | 0.1 (0.0-0.2) |
| < 1 year | 592 (412-777) | 55.5 (38.9-75.0) | 438 (339-654) | 38.1 (29.5-57.0) | 154 (107-207) | 13.4 (9.3-18.0) |
| <5 years | 651 (445-861) | 11.6 (8.6-16.9) | 490 (364-720) | 8.6 (6.5-12.9) | 161 (115-227) | 3.0 (2.1-4.1) |

Abbreviations: CI: confidence intervals.
<sup>a</sup> Medically attended deaths defined as occurring in-hospital.
<sup>b</sup> Non-medically attended deaths defined as occurring out-of-hospital.
<sup>c</sup> Rates expressed per 100,000 population.

