## supplementary methods and tables for "The burden of RSV-associated illness in children aged <5 years, South Africa, 2011 to 2016"

**Supplementary material**

**Methods**

The procedures of this surveillance system have been described in detail in previous publications^1,2,3^. Briefly, dedicated surveillance nurses enrolled influenza-like illness (ILI) and severe acute respiratory illness (SARI) cases Monday through Friday and completed logs of screened patients and total admissions. Records of referral to hospital for ILI cases (ILI cases referred on day of clinic visit are excluded from the analysis) and outpatient care prior to hospitalization were also recorded for ILI and SARI cases, respectively.

Respiratory specimens, nasopharyngeal aspirates or nasopharyngeal swabs, were collected from enrolled patients, placed in universal transport medium (stored at 4-8 °C) and transported to NICD within 72 hours of collection. Specimens were tested for the presence of 10 respiratory viruses (influenza A and B viruses; parainfluenza virus types 1, 2 and 3; respiratory syncytial virus; adenovirus; rhinovirus; human metapneumovirus; and enterovirus) using a multiplex real-time reverse transcription PCR assay^4^.

Methods used to estimate the number and rate (per 100,00 population) of RSV-associated ARI and SARI are described in the following steps:

1. **Estimate the SARI rates in the base provinces (North-West Provinces and KwaZulu-Natal)**

The method for estimation of the rate of SARI is outlined in the formula below:

$$\mathrm{SARI} ratei=\frac{SARI enrolled i*\frac{7}{5}*(\frac{1}{xSARIi})*(\frac{1}{\mathrm{HUS}SARI})}{Popi}$$

Where SARI *ratei* is the age-group hospitalisation rate in age group *i: and SARI enrolled i* the number of SARI cases enrolled in age group *i*: 7/5 is the adjustment for non-enrolment over weekends: *xSARIi* is the proportion of all eligible SRI cases enrolled in each age group *i: HUSSARI* is the Health Care utilisation survey (HUS) derived proportion of SARI cases that sought care at the sentinel hospital^5,6,7–9^: *Pop_i_* is the mid-year population in each age group.

2. **Estimate the SARI rate in other provinces**

The two base province SARI rates (per 100,000) were adjusted for the provincial-level prevalence of risk factors for pneumonia in the other seven provinces obtained from the Demographic Health Survey (DHS)^10,11^. These include HIV infection, exposure to indoor air pollution, crowding, malnutrition, low birth weight and non-exclusive breastfeeding. The relative risks for these risk factors are obtained from published data^10^. The methods is outlined in the formulae below:

$${Adj}_{Y}=\left( 1+\sum_{i} \left( P_{i,Y}-P_{i,B} \right)\times\left( {RR}_{i}-1 \right) \right)$$

Where ${Adj}_{Y}$ is the adjustment factor for the identified risk factors for SARI in province Y; $P_{i,Y}$ is the prevalence of risk factor *i* in province Y from the DHS; $P_{i,B}$is the prevalence of risk factor *i* in the base provinces from the DHS; and ${RR}_{i}$ is the relative risk of SARI due to risk factor *i*.

3. **Estimation of RSV-associated SARI in all provinces**

The provincial rates of RSV-associated hospitalization were calculated by multiplying the provincial SARI hospitalization rates by RSV detection rates (Number of RSV positive/ number tested) obtained from the measured RSV-detection rates at all sentinel surveillance sites (and adjusted by the attributable fraction of RSV)

$RSV{SARI}_{RateH,Y}={SARI}_{RateH,Y} \times{RSVprop}\times{RSV}_{AF}$

Where ${RSVSARI}_{RateH,Y}$ is the RSV-associated SARI hospitalization rate in province *Y* (including the base provinces); ${SARI}_{RateH,Y}$ is the SARI hospitalization rate in province *Y* (including the base provinces); $RSVprop$ is the proportion of SARI cases testing positive for RSV (surveillance data); and ${RSV}_{AF}$ is the attributable fraction of RSV (surveillance data).

4. **Estimation of the number of RSV-associated SARI in all provinces**

We estimated the provincial number of RSV-associated SARI hospitalization by multiplying the provincial RSV-associated SARI rates by the population at risk in each province over the study period^7–9^.

$${RSVSARI}_{NumH,Y}={RSVSARI}_{RateH,Y} \times P{op}_{Y}$$

Where ${RSVSARI}_{NumH,Y}$ is the number of RSV-associated SARI hospitalizations in province *Y* (including the base provinces); ${RSVSARI}_{RateH,Y}$ is the rate of RSV-associated SARI hospitalization in province *Y* (including the base provinces); and $P{op}_{Y}$ is the population in province *Y* ^7–9^.

5. **Non-medically attended**

We used the following 4 steps to estimate the national number and rate (per 100,000 population) of non-medically attended RSV-associated SARI cases. The same methodology as for hospitalized making use of non-medically attended numbers and rates.

a) Estimate the non-medically attended SARI in base province from HUS.

$${SARI}_{RateNH,B}=\left( \frac{{SARI}_{RateH,B}}{{HUS}_{B}} \right)-{SARI}_{RateH,B}$$

Where ${SARI}_{RateNH,B}$ is the rate of non-medically attended SARI in the sentinel hospital catchment population; ${SARI}_{RateH,B}$ is the base rate of hospitalized SARI at the sentinel hospital and ${HUS}_{B}$ is the proportion of all SARI cases that are hospitalized in the base provinces from HUS.

b) Estimate the SARI non-medically attended in other provinces from DHS surveys

$${SARI}_{RateNH,Y}={SARI}_{RateNH,B} \times{Adj}_{Y} \times\frac{{DHS}_{NH,Y}}{{DHS}_{NH,B}}$$

Where ${SARI}_{RateNH,Y}$ is the rate of non-medically attended SARI in province *Y*; ${SARI}_{RateNH,B}$ is the base rate of non-medically attended SARI ; ${DHS}_{NH,Y}$ is the proportion of respiratory cases not seeking care in province *Y* (DHS) ;${Adj}_{Y}$ is the adjustment factor for risk factors of ASRI for province *Y* ; and ${DHS}_{NH,B}$ is the proportion of respiratory cases not seeking care in the base provinces from the DHS.

c) Estimate the non-medically attended RSV-associated SARI by using the RSV detection rate from RSV-associated SARI in the surveillance data/ population in the province

$${RSVSARI}_{RateNH,Y}={SARI}_{RateNH,Y} \times{RSV}\times{RSV}_{AF}$$

Where $RSV{SARI}_{RateNH,Y}$ is the non-medically attended RSV-associated SARI rate in province *Y* (including the base provinces); ${SARI}_{RateNH,Y}$ is the non-medically attended SARI rate in province *Y* (including the base provinces – obtained in Steps 1.b and 2.b); $RSV$ is the proportion of SARI cases testing positive for RSV; and ${RSV}_{AF}$ if the attributable fraction of RSV virus detection to illness obtained from at the same sentinel sites over the same study period [Will add my AF paper once published].

d) Estimate the non-medically attended number of RSV-associated SARI is done by adding all the provincial estimates and dividing by the national population.

$${RSVSARI}_{NumNH,Y}={RSVSARI}_{RateNH,Y} \times P{op}_{Y}$$

Where ${RSVSARI}_{NumNH,Y}$ is the number of non-medically attended RSV-associated SARI cases in province *Y* (including the base provinces); ${RSVSARI}_{RateNH,Y}$ is the non-medically attended RSV-associated SARI rate in province Y (including the base provinces); and $P{op}_{Y}$ is the population in province Y (including the base provinces).

Calculation of the confidence intervals (CI) for medically and non-medically RSV-associated illness is obtained by using bootstrapping resamples over 100 replications for all parameters. The lower limit and upper limit of the CI are the 2.5^th^ and 97.5^th^ percentiles of the estimated values from 1000 resampled datasets.

**Burden of RSV-associated ILI**

ILI surveillance is only conducted at one of several clinics serving each of the base hospital catchment areas, therefore ARI estimates for the base population are derived by a backwards adjustment of the SARI rate at sentinel hospitals, adjusting for ILI cases transferred to hospital and in referrals for other facilities.

As follows:

$${ARI}_{ratei}=\left( {SARI}_{ratei}*Z*X*\frac{Y_{ILIi}}{Y_{SARIi}} \right)*proportion with no fever in age group$$

Where
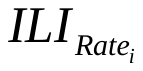
 is the estimated age-specific rate of RSV-associated ILI outpatient consultations (in age group *i*);
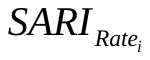
is the age-specific rate of SARI hospitalization (adjusted for non-enrollment and healthcare seeking behavior, as per the calculation for SARI); Z is the proportion of SARI cases that sought outpatient care before hospitalization; X is the ratio of ILI consultations referred to hospital/ the total number of ILI consultations obtained from ILI surveillance;
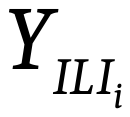
is the proportion of ILI case in age group *i* over the total ILI cases after adjusting for non-enrolment;
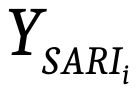
is the proportion of SARI cases in age group I over the total SARI cases after adjusting for non-enrolment

We use ILI at sentinel sites as proxy for the base province, and expand to other provinces and nationally in a similar way to the SARI/RSV-associated SARI estimates, but without adjusting for the SARI risk factors. Finally, ILI estimates are adjusted for the proportion of children who do not present with fever for final acute respiratory illness (ARI) rates^12^.

**Supplementary Table 1: Estimated mean annual number and rates of respiratory syncytial virus-associated mild illness in children aged <5 years, South Africa, 2011-2016.**

| **Age group**  **Months** | **Respiratory syncytial virus-associated mild illness^a^** | | | | | |
| --- | --- | --- | --- | --- | --- | --- |
|  | **Total** | | **Medically-attended** | | **Non-medically-attended** | |
|  | **Number (95% CI)** | **Rate^b^ (95% CI)** | **Number (95% CI)** | **Rate^b^ (95% CI)** | **Number (95% CI)** | **Rate^b^ (95% CI)** |
| <1 | 11543 (0-39394) | 12004.7 (0-40969.2) | 2625 (0-9263) | 2730,2 (0,0-9528,6) | 8918 (0-30231) | 9274,6 (0,0-31440,0) |
| 1 | 10348 (4248-18535) | 10992.8 (4512.8-19689.9) | 2352 (1042-4051) | 2499,0 (1107,3-4302,8) | 7996 (3206-14485) | 8493,7 (3405,5-15386,5) |
| 2 | 17259 (8776-26759) | 18360.5 (9336.3-28466.4) | 3924 (2042-5860) | 4175,0 (2172,5-6233,4) | 13334 (6734-20899) | 14185,5 (7163,6-) 22231,7 |
| 3 | 8737 (4066-14378) | 9308.9 (4332.3-15318.6) | 1986 (932 3148) | 2116,4 (993,0- 3352,9) | 6751 (3134-11230) | 7192,5( 3338,6-11965,1) |
| 4 | 11675 (5769-19465) | 12461.7 (6157.7-20775.9) | 2654 (1337- 4293) | 2833,1 (3352,9-4582,1) | 9021 (4432- 15172) | 9628,6 (4730,8-16193,8) |
| 5 | 5363 (2024-9806) | 5734.8 (2164.3-10486.0) | 1219 (470-2153) | 1303,7 (502,9-2301,1) | 4144 (1554-7654) | 4431,1 (1661,5-8184,3) |
| 6 | 5667 (2399-9640) | 6068.1 (2569.1-10323.2) | 1288 (539-2141) | 1379,3 (577,0-2292,3) | 4379 (1860- 7500) | 4688,9 (1991,6-8031,0) |
| 7 | 6760 (3213-11344) | 7249.8 (3445.5-12166.6) | 1535 (723-2508) | 1646,5 (774,7-2689,4) | 5225 (2490-8837) | 5603,3 (2669,6- 9477,1) |
| 8 | 4343 (1588-7824) | 4664.0 (1705.2- 8402.6) | 988 (365-1766) | 1061,1 (391,2-1896,4) | 3355 (1223-6058) | 3602,8 (1313,5-6505,6) |
| 9 | 7461 (3990-11472) | 8024.9 (4291.7-12338.2) | 1696 (918- 2485) | 1824,5 (986,3-2672,4) | 5765 (3073-8987) | 6200,4 (3304,7-9665,3) |
| 10 | 4241 (1821-7567) | 4571.0 (1962.3-8155.5) | 963 (439-1671) | 1038,4 (472,8-1800,0) | 3278 (1382-5896) | 3532,6 (1489,5-6354,2) |
| 11 | 4506 (1827-7637) | 4873.1 (1976.2-8259.0) | 1024 (421-1685) | 1107,3 (455,0-1821,8) | 3482 (1407-5952) | 3765,9 (1521,2-6436,6) |
| 12-14 | 33621 (19584-50774) | 11935.2 (6952.2-18024.5) | 7645 (4565-11116) | 2713,8 (1581,0-3945,8) | 25976 (15019-39659) | 9221,3 (5331,6- 14078,6) |
| 15-17 | 22157 (11305-36639) | 7865.4 (4013.1-13006.5) | 5036 (2508-7893) | 1787,9 (890,5-2801,5) | 17120 (8796-28746) | 6077,5 (3122,5-10204,5) |
| 18-20 | 23832 (12778-37462) | 8460.4 (4535.9-13298.9) | 5418 (2993-8145) | 1923,5 (1062,1-2891,4) | 18414 (9785-29317) | 6536,9 (3473,5-10407,4) |
| 21-23 | 28903 (12127-49316) | 10260.5 (4305.1 17507.0) | 6570 (2748-10924) | 2332,3 (975,6-3877,8) | 22333 (9379-38392) | 7928,2 (3329,5-13628,9) |
| 24-35 | 32177 (21840-44569) | 2860.9 (1941.8-3962.7) | 7313 (5211-9548) | 650,2 (463,3-848,9) | 24864 (16658-35082) | 2214,5 (1481,0-3119,2) |
| 36-47 | 19704 (12678-27630) | 1799.4 (157.7-2523.1) | 4474 (2960-6031) | 408,6 (270,4-550,7) | 15230 (9717-21599) | 1390,8 (887,3-1972,4) |
| 48-59 | 14404 (8382-22173) | 1316.0 (765.9-2025.9) | 3271 (2804-4789) | 298,9 (174,5-437,5) | 11133 (6472-17384) | 1017,2 (887,3-1588,4) |
| < 1 year | 96783 (39380-181197) | 8615.7 (3505.7-16130.4) | 22002 (9143-40327) | 1958.6 (813.9-3590.0) | 74781 (30237- 140869) | 6657.1 (2691.7-12540.4) |
| <5 years | 264112 (134357- 437187) | 4746.5 (2414.6-7856.6) | 60032 (31176- 96004) | 1078.8 (560.3-1725.4) | 204080 (103181-341183) | 3667.7 (854.4-6132.0) |

Abbreviations: CI: confidence intervals.

^a^ Mild illness defined as not requiring hospitalization; (Obtained from case-based surveillance at 5 facilities and extrapolated nationally).

^b^ Rates expressed per 100,000 population.

**Supplementary Table 2:** Estimated mean annual number and rates of respiratory syncytial virus-associated severe illness (excluding deaths) in children aged <5 years, South Africa, 2011-2016.

| **Age group: months** | **Respiratory syncytial virus-associated severe illness^a^** | | | | | |
| --- | --- | --- | --- | --- | --- | --- |
|  | **Total** | | **Medically attended** | | **Non-medically attended** | |
|  | **Number (95% CI)** | **Rate^b^ (95% CI)** | **Number (95% CI)** | **Rate^b^ (95% CI)** | **Number (95% CI)** | **Rate^b^ (95% CI)** |
| <1 | 14110 (9784-18889) | 14674 (10175-19645) | 6542 (4871-8201) | 6804(5065-8529) | 7568 (4913-10688) | 7871 (5109-11116) |
| 1 | 13872 (11003-17389) | 14736 (11689-18472) | 6431 (5515-7382) | 6832( 5858-7842) | 7441 (5488-10007) | 7905 (5829-10630) |
| 2 | 9848 (7600-12613) | 10477 (8085-13418) | 4566 (3764-5404) | 4857 (4004-5749) | 5282 (3836-7209) | 5619 (4080-7669) |
| 3 | 8721 (6550-11357) | 9292 (6978-12100) | 4043 (3247-4873) | 4308 (3459-5191) | 4678 (3303-6484) | 4984 (3519-6908) |
| 4 | 8263 (5966-11080) | 8820 (6368-11826) | 3831 (2962-4774) | 4089 (3161-5095) | 4432 (3004-6306) | 4731 (3206-6730) |
| 5 | 5251 (3662-7334) | 5615 (3916-7842) | 2435 (1797-3185) | 2604 (1922-3405) | 2816 (1865-4149) | 3011 (1994-4437) |
| 6 | 5721 (4034-7791) | 6126 (4320-8343) | 2652 (1976-3369) | 2840 (2116-3608) | 3069 (2058-4422) | 3287 (2203-4735) |
| 7 | 3862 (2475-5625) | 4142 (2654-6033) | 1790 (1196-2429) | 1920 (1282-2605) | 2072 (1279-3196) | 2222 (1371-3427) |
| 8 | 2595 (1592-3776) | 2789 (1710-4055) | 1203 (750-1645) | 1292 (805-1766) | 1392 (842-2131) | 1495 (904-2287) |
| 9 | 2187 (1211-3493) | 2352 (1302-3757) | 1014 (579-1545) | 1091 (623-1662) | 1173 (632-1948) | 1262 (680-2095) |
| 10 | 2147 (1214-3308) | 2314 (1309-3565) | 996 (584-1467) | 1074 (629-1581) | 1151 (630-1841) | 1241 (679-1984) |
| 11 | 1994 (1102-3180) | 2156 (1192-3439) | 925 (534-1382) | 1000 (578-1494) | 1069 (568-1798) | 1156 (614-1944) |
| 12-14 | 4154 (2855-5955) | 14755 (1014-2114) | 1927 (1397-2564) | 684 (496-910) | 2227 (1458-3391) | 791 (517-1204) |
| 15-17 | 2904 (1861-4200) | 1031 (661-1491) | 1347 (902-1823) | 478 (320-647) | 1557 (959-2377) | 553 (340-844) |
| 18-20 | 2107 (1154-3249) | 748 (410-1153) | 978 (549-1447) | 347 (195-514) | 1129 (605-1802) | 401 (215-640) |
| 21-23 | 2914 (1296-4852) | 1035 (460-1722) | 1352 (624-2181) | 480 (221-774) | 1562 (672-2671) | 555 (239-948) |
| 24-35 | 2831 (1858-4091) | 252 (165-364) | 1313 (899-1792) | 117 (80-159) | 1518 (959-2299) | 135 (85-204) |
| 36-47 | 1355 (620-2326) | 124 (57-212) | 628 (293-1034) | 57 (27-94) | 727 (327-1292) | 66 (30-118) |
| 48-59 | 1384 (643-2343) | 127 (59-214) | 642 (297-1044) | 59 (27-95) | 742 (346-1299) | 68 (32-119) |
| <1y | 78571 (56187-105831) | 6995 (5002-9421) | 36428 (27772-45654) | 3243 (2472-4064) | 42143 (28415-60177) | 3752 (2530-5357) |
| <5y | 96220 (66470-132844) | 1729 (1195-2387) | 44615 (32731-57538) | 802 (58-1034) | 51605 (33739-75306) | 927 (606-1353) |

Abbreviations: CI: confidence intervals.

^a^ Severe illness defined as requiring hospitalization excluding deaths.

^b^ Rates expressed per 100.000 population.

^c^ Obtained from case-based surveillance at 5 facilities and extrapolated nationally.
